# Access to a telehealth falls prevention program: mixed-method analysis from a health equity perspective

**DOI:** 10.64898/2026.01.20.25342838

**Authors:** J O’Neil, N Dionne, Sauvé-Schenk, J Savard

**Author notes:** Corresponding author: Jennifer O’Neil PT, PhD, University of Ottawa, Faculty of Health Sciences, School of rehabilitation sciences 200 Lees Avenue, Ottawa, Ontario.

## Abstract

**Introduction:** Access to falls prevention programs, addressing an important public health concern, is often limited for Francophone and Acadian minority communities. Technology has been used to deliver falls prevention programs, but health equity factors linked to improving its access for all are still poorly understood. This study aimed to document health equity factors influencing access from the perspective of older francophone adults in various communities across Canada.

**Methods:** Guided by the digital health equity framework, we used a sequential explanatory mixed method, including a descriptive sociodemographic questionnaire and semi-structured interviews, to document perspectives on access of persons who had completed different versions of a telehealth falls prevention program.

**Results:** Sixty-one participants from five provinces completed the sociodemographic questionnaire, of which, most participants (n=45) represented perspectives from francophone communities living in a minority situation. Results revealed that 91% of participants had access to a computer or tablet with adequate audio-visual components for telehealth. Twenty of these participants, from four provinces, shared their perspectives. The majority of them reported that falls prevention programs should be offered in their preferred language and with different delivery options that consider the social and safety preferences of participants. Eight recommendations informing policy and implementation emerged.

**Discussion:** Advocacy around digital connectivity is essential to ensure that all Canadians have equitable access to high-speed internet connection and proper infrastructure. Participants also raised the importance of adapting services to their linguistic realities, geographic location, safety and basic needs to optimize the impact of the health services provided to them.

## Context

According to the World Health Organisation (WHO), 684,000 persons are estimated to die following a fall, of which individuals over the age of 60 are most affected(1). Falls can result in physical and emotional trauma and negatively affect quality of life, social participation, and activity of daily life function (dressing, transferring from one position to another, and toileting)(2). Exploring ways to prevent such impact on a populational level, such as targeted implementation of primary falls prevention programs, is crucial. Primary prevention programs aim at decreasing the number of first falls, therefore slowing down health decline linked to injuries following a fall. Primary prevention programs are developed to have an impact on risk factors such as muscle weakness, decreased balance and mobility as well as environmental factors and reduced social participation(3).

To achieve populational-level reach and sustainable implementation, falls prevention programs must consider accessibility and equity-related factors at individual, community, and organisational levels that may influence it. According to the WHO, health equity is defined as “the absence of unfair and avoidable differences in health among groups”(4). Access to health is defined as the fit between a persons’ health needs and the health system(5). Neglecting to consider health equity factors, such as a program cost and geographical location, and the specific needs of a community could negatively impact participation and overall implementation (6,7). At the individual level, the language of program delivery was also reported to influence participation in health programs (8–11). Receiving health services in a preferred language has been associated with client safety and satisfaction (12,13). In Canada, there are two official languages - French and English. The Canadian government recognized official language minority communities as composed of English-Speaking individuals in the Quebec province and the French-speaking individuals outside Quebec. Majority or minority status will affect access to health services in one’s preferred language (14). Community and organization level factors influencing successful implementation of primary falls prevention programs that promote equitable access include community and organisational priorities, financial support, trained program facilitators, and accessible infrastructure (15). However, healthcare program delivery is often limited by the shortage of healthcare providers, and in the case of linguistic minorities, the lack of healthcare providers that can provide services in the patient’s language of preference. This situation leads to reported inequities in care, including difficulty in expressing needs, increased chances of diagnostic errors and poorer outcomes, and delayed consultation (8,11,15,16) More specifically, members of linguistic minorities are less likely to take part in preventive activities not provided in their preferred language (12,15). To improve implementation of accessible primary health prevention programs in Canada that respect principles of equity, programs must be offered in both official languages, English and French. Special attention must be paid to offering programs in French for Francophone living in minority situations, that is, outside the province of Quebec, where French is the majority language.

### The use of Telehealth

Telehealth is a potentially viable solution to reducing geographical barriers. Its use in rehabilitation service delivery offers an alternative option for people to access programs that are not offered in their area(17,18). Specific to falls prevention, telehealth falls prevention group programs can improve access in rural and remote settings and results show promising improvement in gait parameters (19). Other studies have demonstrated the acceptability and usability of virtually supported home-based exercise programs to decrease fall risks in adults living with COPD and Parkinsons (20,21). Some programs have considered the impact of geographical barriers and linguistic barriers in their implementation. For example, *Marche vers le Futur* (MVF) is an effective primary falls prevention program offered for francophone older adults living in a minority situation and available in various telehealth options (15,22).

## Objective and research questions

From a pragmatic and social justice lens, documenting the implementation factors of a falls prevention program that is accessible to underserved older adults and respects equity principles was critical. Our primary objective was therefore to understand the service delivery preferences of older francophone adults who participated in the MVF program and their perspectives on program equity. Our secondary objective was to explore the influence of program cost as an implementation barrier.

## Methodology and approach

We used a sequential explanatory mixed method (23) including a descriptive sociodemographic questionnaire and semi-structured interviews to document the perspectives of persons who had completed different versions of a falls prevention program. Guided by the digital health equity framework (24), we documented perspectives on four levels (i.e., individual, interpersonal, community, societal) and across multiple domains influencing health equity (i.e., biological, digital environment, sociocultural, health system).

### Participants

Participants were recruited from cities in five Canadian provinces (Ontario, Prince-Edward Island, New Brunswick, British Columbia, and Quebec) in communities where previously trained MVF program facilitators were available and who were set up for implementation. In collaboration with local community organizations, new individuals aged 55 and up were recruited to participate in the MVF program via posters, social media, flyers in community centers (e.g., community health centers, francophone community groups, health networks). Since the falls prevention program is a primary prevention intervention, it was key to include people before they are affected by falls was key. Potential participants completed a brief inclusion and exclusion screening tool prior to consenting to the study. They were excluded if they had an unstable cardiac condition, undiagnosed health concerns, if they had fallen more than twice in the last year, or if they needed a mobility aid device for short distances (3 meters). All individuals who completed the program for the first time were invited to share their perspective in a semi-structured interview. This study was part of a larger implementation study approved by the University of Ottawa (H-07-23-9360) and the Bruyere Health Research Institute (M16-23-056) research boards of ethics.

### Sample size

Our descriptive quantitative sample was based on a target sample size of 56 participants which estimated for a larger study which was based on a desired 0.8 power with an alpha of 0.05, and an interrater expected agreement (Kappa) of 0.9 (15% attrition rate)(25). Adequate sample size for qualitative research is still subject to debate. More consistently, sufficient sample size is recognized when information becomes redundant, and code saturation is obtained (26), meaning that no new codes are emerging in data from new interviews. Our sample size was based on pragmatic efforts to have a rich and diverse geographical sample from participants who had completed different versions of the program and suggested saturation can be reached between 5 and 24 semi-structured interviews (26,27).

### Intervention

Marche vers le Futur (MVF) - the intervention in this study - is a twelve-week primary preventive telehealth falls prevention program delivered by trained community health workers. It is currently free of charge for participants; all costs associated with participation, including facility rental, equipment and staff payment, are funded by the various local community organizations implementing the program. It includes a 30-minute pre-program balance and mobility assessment, weekly sessions delivered over 10 weeks, and a 30-minutes post program reassessment. Each of these sessions includes a 15 to 20 minutes guided discussion using educational capsules followed by a 45-60 minutes set of exercises that gradually progresses in difficulty each week. The same program could be offered as an in-person group in a community center with a remote facilitator (i.e., MVF Center), or remotely in each participant’s home with a remote facilitator (i.e., MVF Home). These program delivery options allowed each community health center to choose the option that would facilitate reach of older adults within their respective, often rural or remote areas with limited services for francophones. To our knowledge, this is the only telehealth falls prevention program that offers such a diversity of delivery options and is available in French.

### Data collection

Once electronic consent was obtained, each participant completed a sociodemographic questionnaire (e.g., age, sex, gender, first learned and preferred language, citizenship, geographical location, distance between home and health or community center where program was offered, access to basic technology including internet). Direct and indirect cost variables were also collected to understand economic factors related to program delivery options. Direct costs included distance travelled to a health or community center, transportation cost or access to a transportation mode. Indirect costs included access to high-speed internet, monthly cost of internet, access to a computer or tablet at home to participate in a telehealth program. Once participants had completed the falls prevention program, they were invited to participate remotely via TEAMS or by telephone in a 30-60 minute individual semi-structured interview. During these audio recorded interviews, the same interviewer (JO) and note taker (ND) discussed with each participant their experience of completing a falls prevention program, including program delivery options. Following an interview guide (see appendix), questions focused on factors stemming from the digital health equity framework including socio-economic, cultural, geographical factors as well as perceptions of how health, psychological stressors, access to technology, cost, and experience with telehealth may impact access to a health prevention program.

### Data analysis

We analyzed the socio-demographic data descriptively and used a comparative cost-analysis for data on direct and indirect costs. We compared the data stemming from a francophone community living in a minority situation versus those living in a majority situation. All qualitative data were recorded, transcribed, and analyzed using NVivo14 to facilitate data visualization or organization. Two trained researchers (JO and ND) independently used reflexive thematic analysis following the steps proposed by Braun and Clarck (28,29). Researchers became familiar with the data and completed initial coding. They then conducted a deductive analysis based on domains (i.e., biological, behaviour, environment, sociocultural, health system) and levels (i.e., individual, interpersonal, community, system) drawn from the digital health equity framework (24), which allowed both researchers to generate initial themes. This was followed by an inductive analysis during which the two researchers worked separately to reflect on perspectives and the meaning of their extracts, remaining open to the emergence of new themes. Finally, the two researchers revised together, refined, and contextualized all themes to represent participants’ perspectives.

## Results

### Demographic data and preliminary cost analysis

Of the 67 participants who completed the MVF falls prevention program, 57 participated in the MVF Center and 10 participated in the MVF home version. Out of 67 participants, 61 participants from five provinces completed the sociodemographic questionnaire, 45 representing perspectives from francophone minority communities (Ontario, Prince-Edward Island, New Brunswick, and British Columbia), and 16 participants representing perspectives from francophone majority communities (Quebec). The majority of participants were women (n=50) born in Canada (n=57). Out of 61 participants living in francophone communities; 58 reported French as their mother tongue and 59 as their preferred official language for services. Most participants (n=45) lived in a 10km radius of where the program was delivered (figure 1). Only seven participants from rural areas of Quebec and five participants from minority francophone communities lived further than 10km from where the falls prevention program was offered. Only two out of 61 participants documented challenges with transportation to reach the center where the program was offered. The majority of participants (n=51) reported that the cost associated with travel to the program was less than 5 Canadian dollars ($ CAD) per session (60$ CAD in total for 12 sessions).

**Figure 1.**
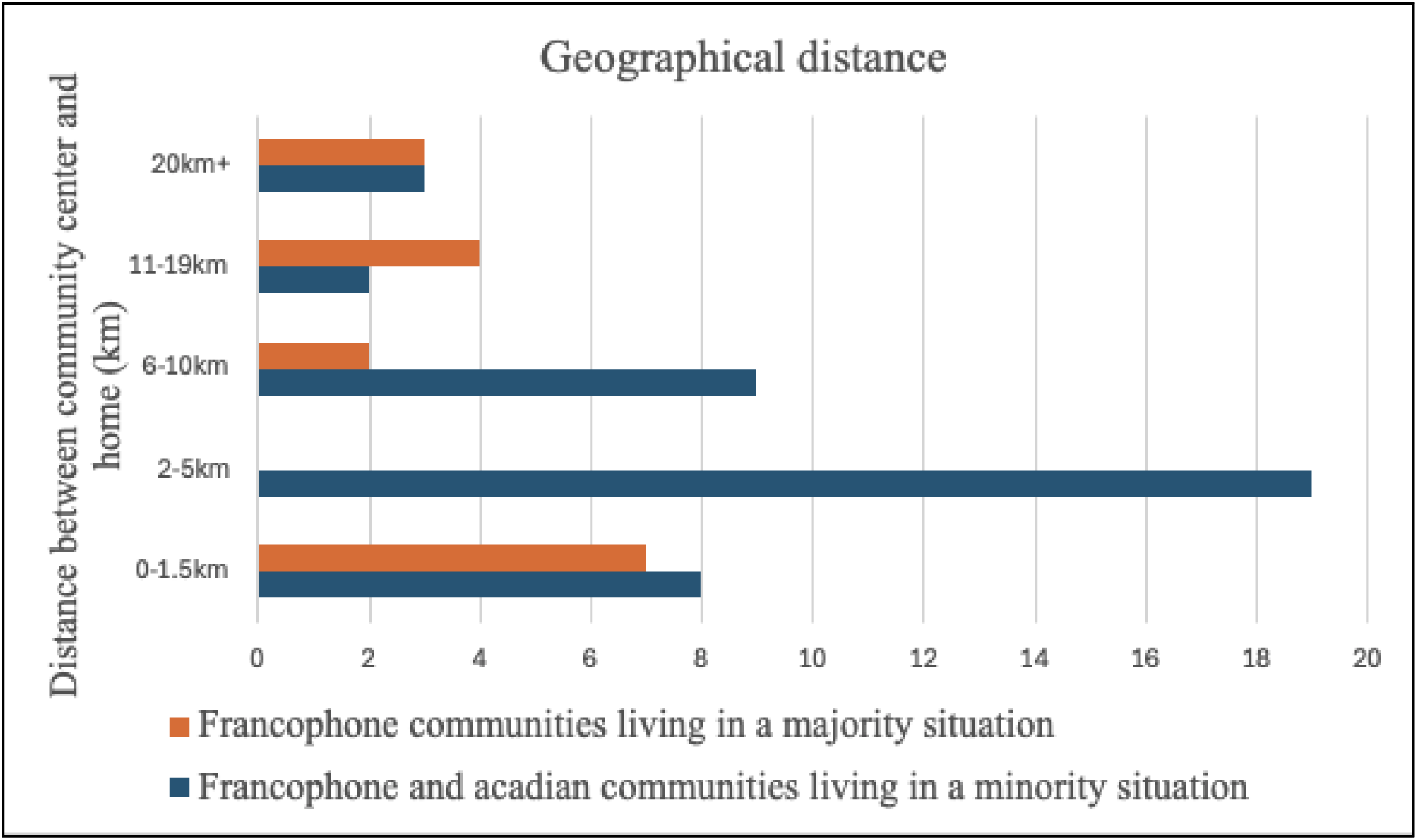
The distance travelled by participants to a community center to participate in a falls prevention program. Comparing the distance reported by francophone minority versus majority communities.

Participant reported indirect technology costs revealed that 91% (59/61) of those who completed the sociodemographic questionnaire had access to a computer or tablet with adequate audio-visual components for telehealth and that 98% (60/61) of participants declared that they had access to high-speed internet (i.e., 50 Mbps for downloads and 10 Mbps for uploads) required to conduct a telehealth session in their home (30). However, the monthly cost of internet varied greatly and revealed geographical disparities. In general, internet cost was higher for francophone minority communities (41/61) compared to francophone majority communities (16/61), which reflect differences in rurality (figure 2). A noticeable higher cost was documented for rural and remote francophone communities in New-Brunswick.

**Figure 2.**
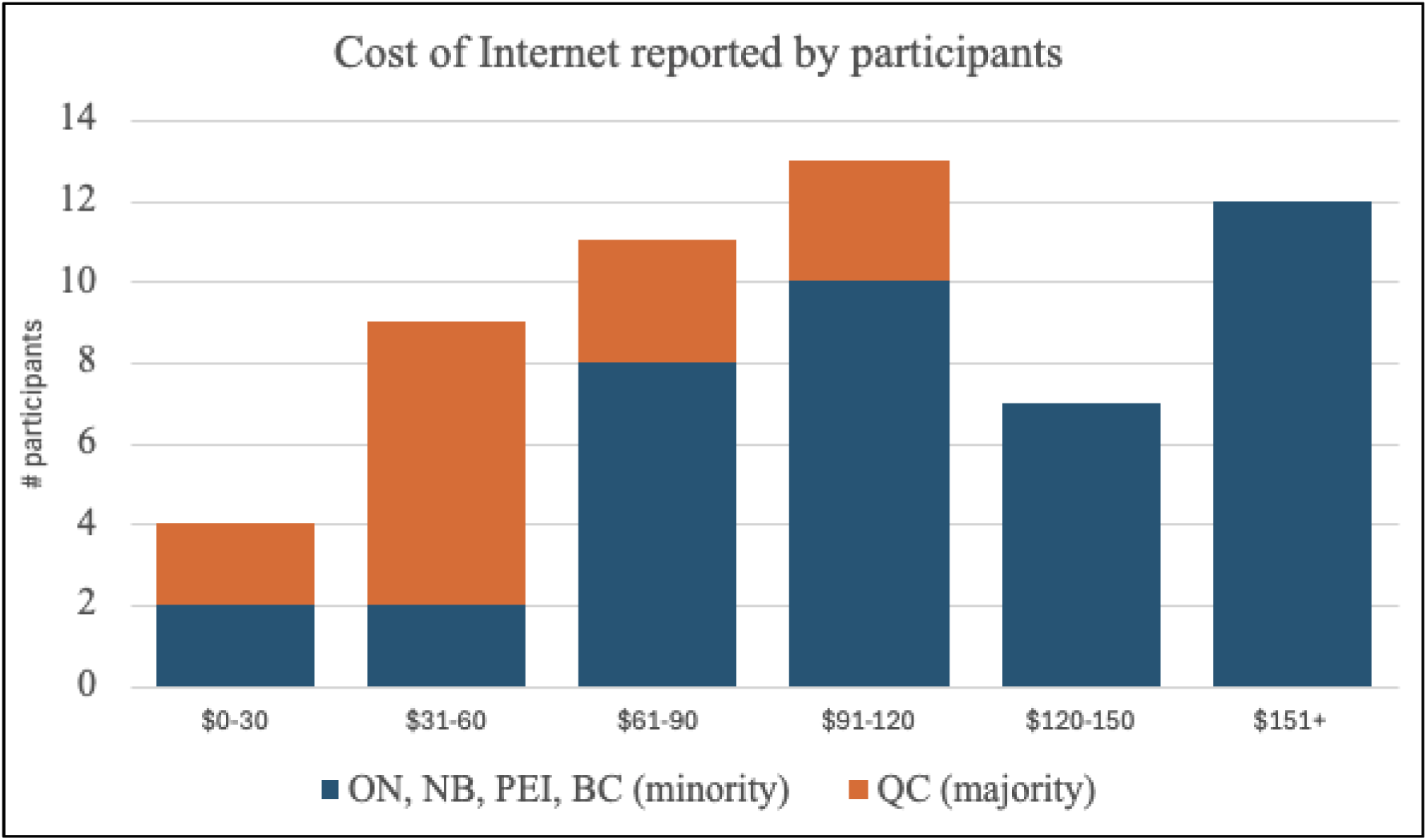
Cost comparison between francophone minority and majority regions

### Participants’ perspectives on equitable access

Twenty participants shared their perspectives on access to a community-based falls prevention program during semi-structured interviews. Participants lived in various locations including rural Quebec (n=3), New-Brunswick (n=9), Prince-Edward Island (n=3), and Ontario (n=5). Based on the digital health equity framework (24), seven themes emerged including three at the individual, two at the interpersonal and community, and two at the societal levels respectively. For each theme, influential implementation concepts for program delivery and program access for older francophone adults were identified. These findings are illustrated with quotes, which were translated from French to English.

## Individual level

### Theme 1: I would not have participated if the program was not offered in French

The majority of participants reported that the program being offered in French, their preferred language, was extremely important to them.

> *“Any source of information or service is good, especially when available in French! In rural and remote areas, we need it.” (P14)*

Without an option to participate in French, participation would be negatively impacted. However, the preferred language did not influence the choice of program delivery option (i.e., in-person, MVF Center or MVF Home). Participants saw the language in which the service was offered either as a participation barrier (e.g., English) or opportunity (e.g., French) first, prior to focusing on where and how the service was being delivered.

> *“If it was not offered in French, I would not have participated. You do not always understand everything if it’s not in your first language, no matter how its delivered.” (P2)*

### Theme 2: I would not have participated if I could not afford it

Participants raised the potential impact of cost and its relationship with how a person chooses to access a falls prevention program. The main cost variables discussed were technology and transportation.

> *“The cost is higher in-person/at the community center, but I prefer this option [because I could afford it][…] If you do not have money for a computer, you cannot participate. If you live far [from the community center], you cannot afford a car to travel.” (P2)*

When we asked them to reflect on affordability and what an acceptable fee could be for a twelve-week falls prevention program similar to the one they participated in, they suggested between 10$ and 200$ per program. Most voiced that they would be comfortable paying 5$ to 10$ per session but acknowledged that the program being free allowed for better access for all.

> *“It’s difficult to mobilize people here, but since the program was free, people were really interested in our region. At 10$ or less pers session, I think people would still join.” (P20)*

### Theme 3: My emotional and physical safety will influence my decisions related to how I access

Participants described the importance of considering the mobility level, functional capacity, and how emotional and physical safety may influence the type of program delivery for individuals aged 55 and up. First, physical accessibility was raised as a factor to consider when offering a falls prevention program in a community center.

> *“People who have less mobility could have difficulty accessing the center in-person. Maybe these people could participate remotely from their homes.” (P13)*

Second, emotional security was raised as being fundamental, especially for people who experience anxiety or mental health issues.

> *“For example, anxiety could lead to people preferring the remote program at home […] If a person experiences depression or has caregiving duties, leaving the house might be too much effort. For others, it could be nice to get out of the house, meet a group. We had a lot of compassion for each other. It depends on the needs of each person.” (P13)*

Finally, participants also shared their concerns with physical safety in areas where the program is offered. They talked about considering the time of day when the sessions were offered as well as social realities which could impact safety in certain locations.

> *“There are a lot of violence and drug problems, people will only go out during the day. The security of participants is important.” (P9)*
>
> *“In the evening, a remote program at home would be better for safety.” (P12)*

## Interpersonal and community levels

### Theme 4: I need social interactions

Gathering socially during falls prevention program sessions was described as essential for the wellbeing of participants. The informal interactions, group discussions before, during, and after the program allowed participants to develop interpersonal relationships. Participants reported that these social interactions were a deciding factor in their choice to access the program in a community center versus the option at home.

> *“I live alone so I need social time. I love the interaction between participants. I find it hard to do it on my own, I need to be motivated by others.” (P14)*
>
> *“Personally, I like it better in a group. There is more contact, we understand better and we help each other. It’s more intimate that way. I live alone so I like the bonds that it creates” (P15)*
>
> *“I really would have liked to participate in-person, but it wasn’t possible so I’m happy I had the possibility to do it at home.” (P12)*

### Theme 5: I appreciate the telehealth option for rural and remote areas

Participants appreciated having a telehealth option for rural and remote areas where the infrastructure or access to transportation may not support in-person delivery or MVF Center delivery. Having different delivery options, such as telehealth, made it possible to adapt programs to regional realities. Many participants voiced that having access to programs offered in French was often limited by geographical location, with prevention services in rural and remote areas being fewer than in urban areas.

> *“Rural areas are abandoned in favor of big cities, it does not matter which city or province, it has always been like this. Healthcare is inaccessible.” (P4)*

Other participants indicated that the distance between their home and the community center where services are provided was a limiting factor for access. Some participants suggested that four to five kilometers between the two was acceptable, whereas greater distances would influence their choice of program type (e.g., community center versus home-based). Specifically, they shared that 15km would be possible depending on the time of the day but anything greater than 30km would lead to accessing a program remotely, delivered at home.

> *“The easiest option for me is by video [remote at home]. It would be 45 minutes from my house to go to a center. Public transport is not reliable.” (P17)*

## Societal and health system level

### Theme 6: I suggest integrating falls prevention with other community health services

When reflecting on healthy aging, participants considered falls prevention as a basic need. They also suggested that balancing basic needs, such as housing, adequate nutrition and health prevention services, should be carefully considered. To this end, they proposed that community solutions could facilitate access to some of these basic needs simultaneously.

> *“Unless someone is really struggling financially [basic needs]… that could maybe have an influence. Maybe volunteers could be available to drive people [to the community center so they have access]” (P1)*
>
> *“To help [with basic needs], we could collaborate or partner with a soup-kitchen that could offer nutritional snacks at the end of a falls prevention program.” (P4)*

### Theme 7: Telehealth should be considered as a solution for certain health and prevention services, under certain conditions, and for certain individuals

The majority of the participants shared that telehealth has a place in the Canadian health system. They provided examples of appropriate services such as renewing prescriptions or asking questions to family doctors. They highlighted that the use of videoconference (such as experienced during the telehealth falls program) was preferred over a phone call. However, they suggested that digital literacy training be adapted to francophones living in a minority situation to facilitate its use.

> *“Of course, having access to telehealth [medical services] helps. It will free up emergency departments in hospitals, but I am not sure if there are many people who feel comfortable calling or using the internet to receive treatments compared to being in-person. Personally, I would like videoconferencing much more than phone calls, but my doctor has never offered this option. I need to see the person. It might take a little longer, but I believe that by seeing the patient, the doctor could identify issues that would not be identified over the phone.” (P8)*

## Discussion

The use of telehealth is growing even if some health professionals remain uncertain about older adults’ ability to manage health technology(31,32). To date, few studies have investigated how remote delivery options may impact access to health prevention programs for older francophone adults. Specific to falls prevention, our results align with current trends to provide sensitive, responsive, and targeted falls prevention programs (33). Our study offers important recommendations around the need for alternative telehealth options based on geographical context, costs, and specific health equity considerations. Having different delivery options could provide an added value from a health equity and participation perspective.

When comparing the use of telehealth in a community center (in-person participants with remote facilitator) versus telehealth at home, participants preferred a community center version. The social aspect linked to this version was indispensable. Providing opportunities for social interactions is suggested to facilitate engagement, which is essential to participation(34). It has also been shown to benefit adherence to telehealth services(35). Social support and friendships developed between participants could contribute to reducing isolation(6,34). Since isolation is associated with a fear of falling(36), which is a recognized as risk factor for falls, promoting social interactions between participants could contribute to reducing both fear of falling and secondary falls. Interpersonal relationships may also positively influence participants to overcome barriers such as lack of public transport and cost associated with travel by creating carpooling networks between participants. Despite a preference for remote delivery in a community center, offering an alternative option that was accessible from home was welcomed for individuals with reduced mobility, limited transportation or those who did not feel safe within their community. Perspectives seemed to vary depending on the delivery mode in which participants completed the program. Individuals who participated in the in-person version tended to highlight the importance of social interactions whereas individuals who had completed the remote version emphasized ease of access and absence of travel costs. Considering these factors in the implementation of programs is essential to offer sensitive, responsive, and equitable services.

Our study allowed us to compare participants’ perspectives on access between francophone living in a minority situation and those living in a majority situation. French is the first official language spoken by 22% of the Canadian population; these people account for 84% of the population in Quebec (majority community), 30% in New-Brunswick, and 26% of other provinces and territories (minority communities) combined(37). Linguistic discordance is an additional barrier to access often combined with geographical barriers(38). While innovation such as telehealth is documented as being a factor associated with successful implementation of falls prevention programs(6), considering linguistic concordance is crucial to ensure that programs are equitable and adapted to the needs of each community. Findings from our study revealed that for francophones in minority or majority situations, having the choice to participate in a falls prevention program in-person or remotely, in a community health center or at home was appreciated as long as the delivery was provided in French by a trained facilitator.

Our preliminary cost analysis showed that cost of internet was an important element to consider. Indirect cost linked to accessing high-speed internet varied greatly across geographical areas. In 2023, Statistic Canada reported that only 95,6% of household in New-Brunswick had the possibility to connect to high-speed internet compared to 100% in Quebec(39). Our study also documented a provincial disparity for participants living in New-Brunswick who access high-speed internet at a higher cost. Given that a larger proportion of Francophones in minority situation reside in rural areas (40,41), this disparity in internet access will have a greater impact on these communities. These results reinforce the literature on the impacts of geographical region, socio-economic status, linguistic preference, and proper infrastructure and information on implementation of health programs(40,42). Advocacy around digital connectivity is essential to ensure that all Canadians have equitable access to high-speed internet connection and proper infrastructure.

Finally, two important elements emerged during our semi-structured interviews. First, older francophone adults voiced the importance of accessing programs that considered emotional and physical safety. Participants who feared for their safety in certain communities or neighborhoods shared that they appreciated having the option to participate from their homes. This lived experience aligns with results from a scoping review by Kepper et al., suggesting that social and environmental characteristics putting people in a non-safe space can negatively influence physical activity(43). Offering a telehealth falls prevention program within a safe space could positively impact the level of physical activity. Second, discussions from participants around basic needs such as food security, have highlighted important considerations in the provision of prevention programs. Food security can significantly impact the health and wellbeing of older adults(44), and contribute to fragility(45). Since fragility can increase the risks of falls, it would be pertinent to integrate food security or a nutritional element to the development of future community falls prevention programs.

### Limitations and future opportunities

Although participants were spread across five provinces for the socio-demographic and cost analysis, the interviews were only representative of five communities across four provinces. The saturation of codes of the interview analysis increases the credibility of our qualitative findings for population similar to our sample. However, our sample was not representative of all geographical areas and lacked diversity from a sex and gender lens. It is therefore possible that implementation realities may differ in the Canadian northern territories or Canadian prairies, since our study did not include any participants from these regions. Older francophone males were also underrepresented, which was not surprising since it is well known in the literature that females participate in falls prevention program more than males(46,47). When asked about male participation, participants mostly shared preconceived ideas around group exercise, often seen as targeting females. It would be critical to further understand the needs and perspectives of males to improve male participation in falls prevention. Despite these limitations, sociodemographic data and health equity perspectives across different provinces provided a holistic and varied view around access to a telehealth falls prevention program.

All participants in this study completed the MVF program which could potentially bias the experiences of individuals based on the program content specific to MVF. Exploring the experiences of individuals participating in different fall prevention programs could perhaps provide a broader understanding of participants’ perspectives regarding access and falls prevention program implementation. Following health equity principles, aligning delivery options with the needs and resources of each community was key. Unfortunately, due to limited resources in each community organisation, participants did not have the opportunity to choose their preferred delivery option, which is acknowledge as a limitation. To mitigate this potential bias, during the interviews we provided participants with detailed examples of the delivery option they had not experienced.

### Health-equity and access considerations: 8 recommendations

Eight recommendations, based on perspectives of participants from four provinces, can support advocacy actions for political decision-makers, guide program delivery choices of health network managers, and inform the development and implementation of new falls prevention programs or equitable telehealth services.

1. Falls prevention should be considered as a basic need.
2. Healthcare should be provided in the person’s preferred language.
3. Socio-economic status should be considered when providing telehealth.
4. Emotional and physical safety should be considered when designing telehealth prevention programs.
5. Telehealth prevention programs must offer opportunities for social interaction.
6. The implementation of telehealth programs in rural and remote areas must be adapted to the available infrastructure and available public transport.
7. Telehealth programs must align with an individual’s needs (specific to conditions) and preferences (for certain types of healthcare).
8. Specific research on sex and gender is needed to better understand how to reach underrepresented groups in falls prevention programs.

## Conclusion

Results from our study allowed us to document lived experiences and perspectives from francophone older adults on health equity and access to a telehealth falls prevention program. It provided the opportunity to compare different delivery options as well as perspectives across geographical areas. Findings demonstrate the importance of adapting the implementation and delivery based on the needs and realities of each francophone community by considering the intersections between basic needs, geography, linguistic concordance, socio-economic status, and safety. Approaching falls prevention or telehealth service implementation from a holistic and health equity lens could positively impact the number of falls in Canada and improve access to preventative services for older adults, including francophones living in a minority situation.

## Acknowledgments section

We acknowledge our community partners and all participants who provided their perspectives which informed our results.

## Ethical approval and informed consent statements

This study was approved by the University of Ottawa (H-07-23-9360) and the Bruyere Health Research Institute (M16-23-056) research boards of ethics.

## Data availability statement

Data collection was completed in French and is available upon request since translation is necessary.

