## Supplemental material for "Access to a telehealth falls prevention program: mixed-method analysis from a health equity perspective"

**Équité et l’efficacité de différentes versions de prestation du programme de télésanté de prévention des chutes Marche vers le Futur**
**Guide d’entrevue**

***English Translation of this interview guide is provided by authors**

| **Au niveau personnel (Personal level)** | | |
|---|---|---|
| **Contexte selon cadre conceptuel** | **Questions et sous-questions (original version)** | **English Translation** |
| Accès et utilisation des ressources de promotion de santé<br><br>(Access to and use of health promotion resources) | Q1. Quelles sont **vos perspectives ou votre point de vue face aux différentes façons** de participer? Dans le programme? | Q1. What are your perspectives or your point of view regarding the different ways of participating in the program? |
| Socio-économique, culturel and politique<br><br>(Socioeconomic, cultural and political) | Q2. Pensez-vous que votre **situation financière/votre revenu** à un impact sur votre choix de participer au programme en personne ou de façon virtuelle?<br><br>Ex. Le fait de vivre dans une situation de faible ou haut revenu affecte t'il votre choix?<br><br>Si oui, comment? Si non, comment? | Q2. Do you think your financial situation/income has an impact on your choice to participate in the program in person or virtually/from home?<br><br>Example: Does living in a low-income or high-income situation affect your choice? |
| Stratification, situation et circonstance sociale<br><br>(Stratification, situation and social circumstances) | Q3. Pensez-vous que votre **langue maternelle ou de préférence à** un impact sur votre choix de participer au programme en personne en salle ou de chez vous virtuellement?<br><br>Ex. MVF Salle, MVF domicile, présentiel<br><br>Si oui, comment? Si non, comment? | Q3. Do you think your first language or preferred language has an impact on your choice to participate in the program in person, in the gym, or virtually/from home? |
| Circonstances matérielles et besoins<br><br>(Material circumstances and needs) | Q4 Pensez-vous que vous avez des **priorités de base qui influencent votre choix?** Par priorités de bases, je veux dire vous loger, vous nourrir, etc. | Q4. Do you think you have basic priorities that influence your choice? By priorities, I mean housing, food, etc. |

| | | |
|---|---|---|
| | Si oui, comment? Si non, comment? | |
| Biologie<br><br>(Biology) | Q5. Est-ce que votre **sexe ou identité de genre affecte votre choix**? Par exemple, être ou vous identifier comme un homme, une femme ou autrement? | Q5. Does your sex or gender identity affect your choice? For example, being or identifying as a man, a woman, or otherwise? |
| Stresseurs psychosociaux et Résilience et résponse au stress<br><br>(Psychosocial stressors and resilience/stress responses) | Q6. Avez-vous vécu **des stresseurs psychologiques** qui pourraient avoir un impact sur votre choix de faire un programme en salle, à domicile, à distance ou en présentiel?<br><br>Par exemple, vous n'aimez pas conduire l'hiver... ou vous êtes souvent seule donc préférez être en salle... | Q6. Have you experienced psychological stressors that could affect your choice to do a gym-based, at-home, remote, or in-person program? |

| **Au niveau communautaire ou sociétal (Community or Societal Level)** | | |
|---|---|---|
| **Context selon cadre conceptuel** | **Questions et sous-questions** | **English Translation** |

| | | |
|---|---|---|
| Environnement<br><br>(Environment) | Q7. Pensez-vous que **l'endroit où vous vivez a un impact sur** votre choix de faire un programme en salle, à domicile, à distance ou en présentiel?<br><br>Ex. Le fait de vivre dans une région rurale ou urbaine affecte-t-il votre choix?<br><br>Q8. Pensez-vous que **l'infrastructure,** par exemple, l'**accès à l'internet, une salle communautaire accessible à un** impact sur votre choix de faire un programme en salle, à domicile, à distance ou en présentiel? | Q7. Do you think the place where you live has an impact on your choice to do a gym-based, at-home, remote, or in-person program?<br>Example: Does living in a rural or urban area affect your choice?<br><br>Q8. Do you think infrastructure, for example access to the internet or an accessible community center, has an impact on your choice to do a gym-based, at-home, remote, or in-person program? |
| Environnent digital<br><br>(Digital environment) | Q9. Pensez-vous que votre **confiance ou aise avec l'utilisation de la technologie à un** impact sur votre choix de faire un programme en salle, à domicile, à distance ou en présentiel?<br><br>Q10. Pensez-vous de **l'utilisation de la santé digitale et la télésanté peut améliorer l'accès aux services de santé au Canada**?<br><br>Est-ce qu'avoir différentes options de programme incluant la télésanté est important pour vous? | Q9. Do you think your confidence or comfort with using technology has an impact on your choice to do a gym-based, at-home, remote, or in-person program?<br><br>Q10. Do you think the use of digital health and telehealth can improve access to health services in Canada?<br>Is having different program options, including telehealth, important to you? |
